# A flexible, pan-species, multi-antigen platform for the detection and monitoring of SARS-CoV-2-specific antibody responses

**DOI:** 10.1101/2021.01.20.21249279

**Authors:** Huifeng Shen, David Forgacs, Digantkumar Chapla, Kelley W. Moremen, Lance Wells, Sarah A. Hamer, Stephen M. Tompkins, Ted M. Ross, Nadine Rouphael, Srilatha Edupuganti, Matthew H. Collins, Rick L. Tarleton

**Affiliations:** Center for Tropical and Emerging Global Diseases, University of Georgia, Athens GA, USA; Center for Vaccines and Immunology, University of Georgia, Athens GA, USA; Complex Carbohydrate Research Center, University of Georgia, Athens GA, USA; Department of Biochemistry, University of Georgia, Athens GA, USA; College of Veterinary Medicine and Biomedical Sciences, Texas A&M University, College Station TX, USA; Department of Infectious Diseases, University of Georgia, Athens GA, USA; Hope Clinic of the Emory Vaccine Center, Division of Infectious Diseases, Department of Medicine, School of Medicine, Emory University, Decatur, Georgia, USA; Department of Cellular Biology, University of Georgia, Athens GA, USA

**Author notes:** Corresponding Author: Rick L. Tarleton, Center for Tropical and Emerging Global Diseases, Coverdell Center for Biomedical Research, 500 D.W. Brooks Dr, University of Georgia, Athens, GA 30602. The authors have declared that no conflicts of interest exists.

## Abstract

The SARS-CoV-2 pandemic and the vaccination effort that is ongoing has created an unmet need for accessible, affordable, flexible and precise platforms for monitoring the induction, specificity and maintenance of virus-specific immune responses. Herein we validate a multiplex (Luminex-based) assay capable of detecting SARS-CoV-2-specific antibodies irrespective of host species, antibody isotype, and specimen type (e.g. plasma, serum, saliva or blood spots). The well-established precision of Luminex-based assays provides the ability to follow changes in antibody levels over time to many antigens, including multiple permutations of the most common SARS-CoV-2 antigens. This platform can easily measure antibodies known to correlate with neutralization activity as well as multiple non-SARS-CoV-2 antigens such as vaccines (*e*.*g*. Tetanus toxoid) or those from frequently encountered agents (influenza), which serve as stable reference points for quantifying the changing SARS-specific responses. All of the antigens utilized in our study can be made in-house, many in *E. coli* using readily available plasmids. Commercially sourced antigens may also be incorporated and newly available antigen variants can be rapidly produced and integrated, making the platform adaptable to the evolving viral strains in this pandemic.

**Brief Summary:** A multi-antigen assay for monitoring SARS-CoV-2-specific antibodies irrespective of host species, antibody isotype, and specimen type was developed.

## Introduction

Since the novel coronavirus SARS-CoV-2, the virus that causes COVID-19, emerged as a worldwide threat in late 2019, a wide variety of tools to detect infection and immune responses to SARS-CoV-2 have been developed. Laboratory tests that detect viral components, specifically nucleic acids via RT-PCR or protein antigens have been most commonly used to diagnose cases of COVID-19. While serological tests have not been widely used in the clinical setting, such assays may have considerable benefit in understanding infection history (for example, in asymptomatic infections where virus detection assays were not conducted), tracking the antigen specificity and longevity of antibody responses over time, and in defining correlates of protection against infection or reinfection. Serological assays are also likely to have great utility in detecting and tracking re-exposure to viral antigens, either as a result of reinfection or vaccination.

The goal of this project was to develop a robust, inexpensive and adaptable serological tool for detecting SARS-CoV-2-specific antibodies and monitoring these responses over time. Our previous experience using the Luminex multiplex bead array platform to detect changes in antibody responses following therapeutic treatment of subjects infected with *Trypanosoma cruzi*, the agent of human Chagas disease, demonstrated the species- and antigen-flexibility, as well as the quantitative precision of this platform (1-7).

Herein we demonstrate the ability to detect and accurately track isotype-specific serological responses to multiple SARS-CoV-2 proteins and protein fragments as well as other antigens, in a variety of species and using multiple sample types. The target antigens used in the assay can be relatively easily produced in-house or are available commercially, making the assay less expensive and readily modifiable to suit specific research questions.

## Results

SARS-CoV-2 Spike and the receptor-binding domain (RBD) fragment of Spike produced in mammalian cells are commonly used for measurement of antibodies to SARS-CoV-2. As protein production in *E. coli* is often cheaper and easier, we initially made codon-optimized Spike, RBD and nucleocapsid proteins (NP) in *E. coli* and compared these to mammalian-expressed Spike and RBD proteins. While the Spike and RBD proteins produced in mammalian cells provided strong specific detection of IgG in sera from virus-positive subjects, the *E. coli*-produced proteins did not (Fig. 1). However, the NP produced in *E. coli* detected strong IgG levels in virus-positive subjects with modest background in the uninfected controls.

**Figure 1.**
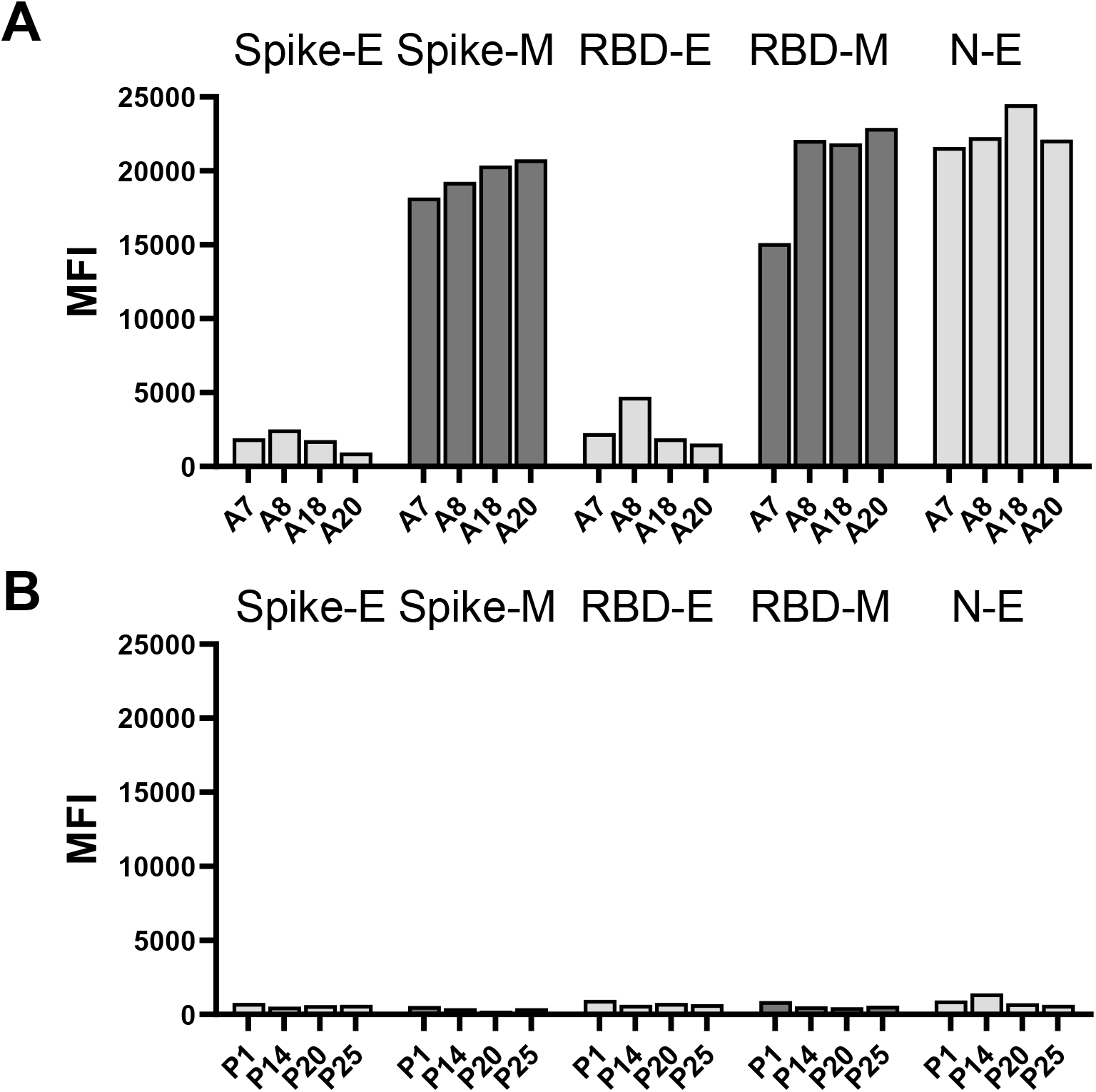
IgG in SARS-CoV-2 Spike protein ELISA-positive sera, selectively recognize Spike and RBD proteins produced in mammalian cell culture but not that produced in *E. coli* (A) while sera from SARS-CoV-2 ELISA-negative subjects recognize none of the SARS-CoV-2 antigens irrespective of source (B). SARS-CoV-2 NP produced in *E. coli* binds antibodies in the SARS-CoV-2 reactive sera (A).

A number of SARS-CoV-2 proteins other than the surface Spike/RBD and nucleoprotein have been noted to be targets of antibody responses in COVID patients, in particular OFR3b and ORF8 (Hachim, et al., 2020). *E. coli*-produced versions of ORF3b and ORF8 exhibited limited detection of antibodies in cohort 1 subjects, including those with strong Spike- and NP-specific IgG and/or IgM responses (Fig. 2a-c).

**Figure 2.**
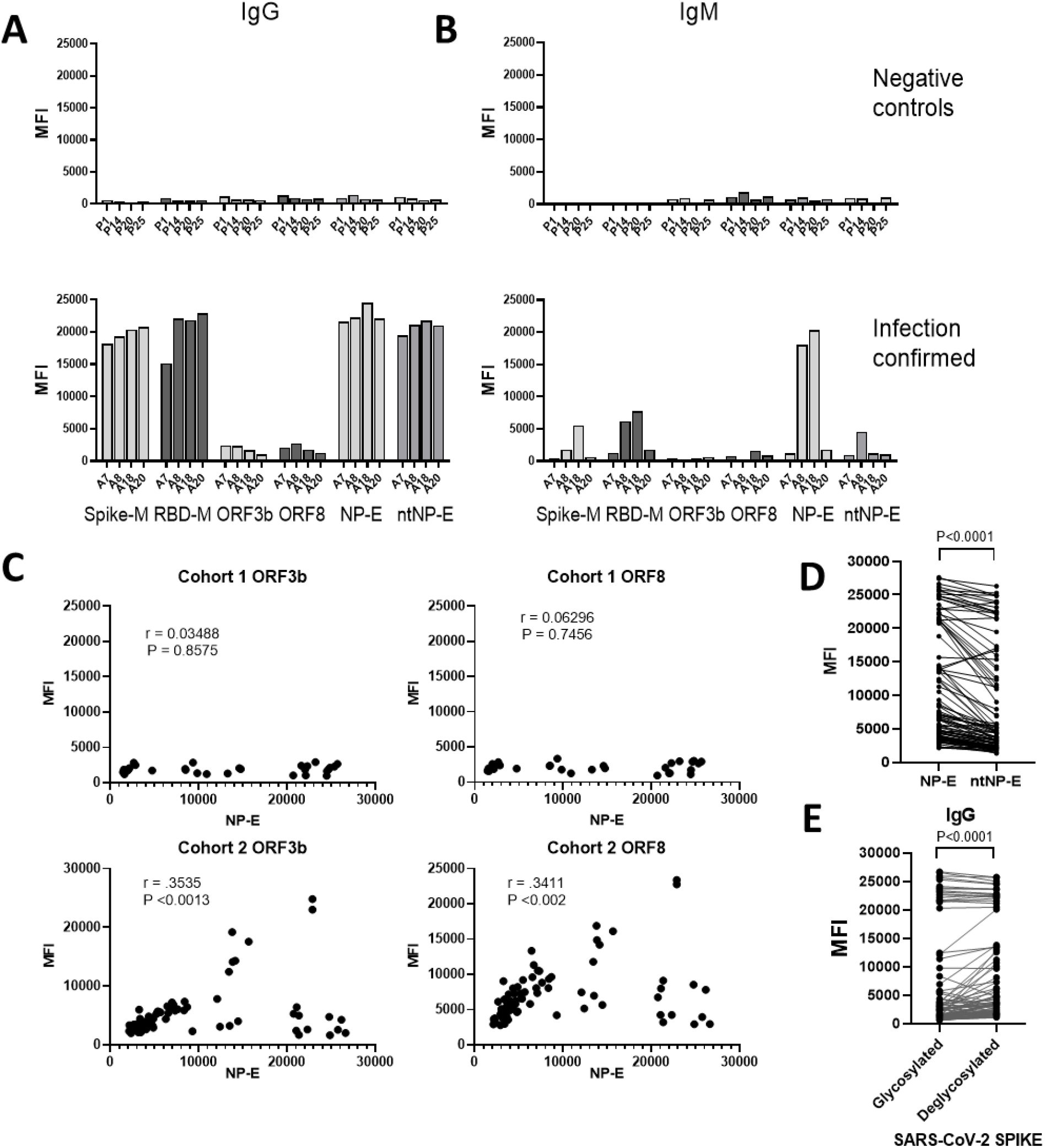
Both IgG (A) and IgM (B) responses to multiple SARS-CoV-2 antigens are evident in selected Cohort 1 but not control subjects. IgG responses to ORF3b and ORF8 are undetectable in Cohort 1 subjects, irrespective of responses to NP-E, but correlate with the NP-E responses in Cohort 2 subjects (C). Relative to the full-length NP-E, the n-terminal fragment of NP-E (ntNP-E) detects significantly less IgG in sera from Cohort 2 and 3 subjects (D). Deglycosylation of Spike-M enhances detection of Spike-specific IgG (E). Matched samples from each subject are connected (D and E). Pearson correlation coefficient and two-tailed P value shown in C. For D and E, P value determined from two-tailed paired T test.

However, in cohort 2, antibodies to both ORF3b and ORF8 were more evident and showed a positive correlation with anti-NP antibodies (Fig. 2c). The n-terminal region of NP (AA residues 47-173; ntNP) has been reported to exhibit lower background responses in nonCOVID-19 subjects, relative to the full-length NP (Phan et al, 2020), but we found that the truncated NP also detected substantially less IgG in the majority of subjects, in some cases nearly totally extinguishing the anti-NP signal detected (Fig 2d).

Considering the heavy glycosylation of the Spike trimer and the failure of Spike produced in *E. coli* to detect anti-Spike antibodies (Fig 1), we asked if glycan groups on the Spike protein might contribute to the detection of anti-Spike antibodies. Somewhat surprisingly, we found that deglycosylated recombinant Spike-M protein bound significantly more antibody in the majority of subjects with low to moderate anti-Spike antibody levels (Fig. 2e).

A major advantage of the Luminex system relative to standard ELISAs or other variants (e.g. Luciferase Immunoprecipitation System (8)) is the ease with which new or modified antigens can be incorporated into the platform without a requirement for increased sample consumption. Anticipating the need to also assess other respiratory infections, as well as the overall immune status of subjects with possible SARS-CoV-2 infections, we incorporated influenza nucleoproteins (A and B), produced in *E. coli*, as well as commercially sourced tetanus toxoid (TT; Sigma Chemicals) Fig. 3 illustrates the wide variation in isotype-specific responses to SARS-CoV-2 antigens in the cohort 2 in contrast to their more uniform and strong Flu-B NP and TT IgG responses. This result corresponds well to the expected pattern of immunity in this cohort: Healthcare workers experienced diverse SARS-CoV-2 exposure conditions at work and in the community, particularly early in the pandemic, and the vast majority receive annual influenza vaccination.

**Figure 3.**
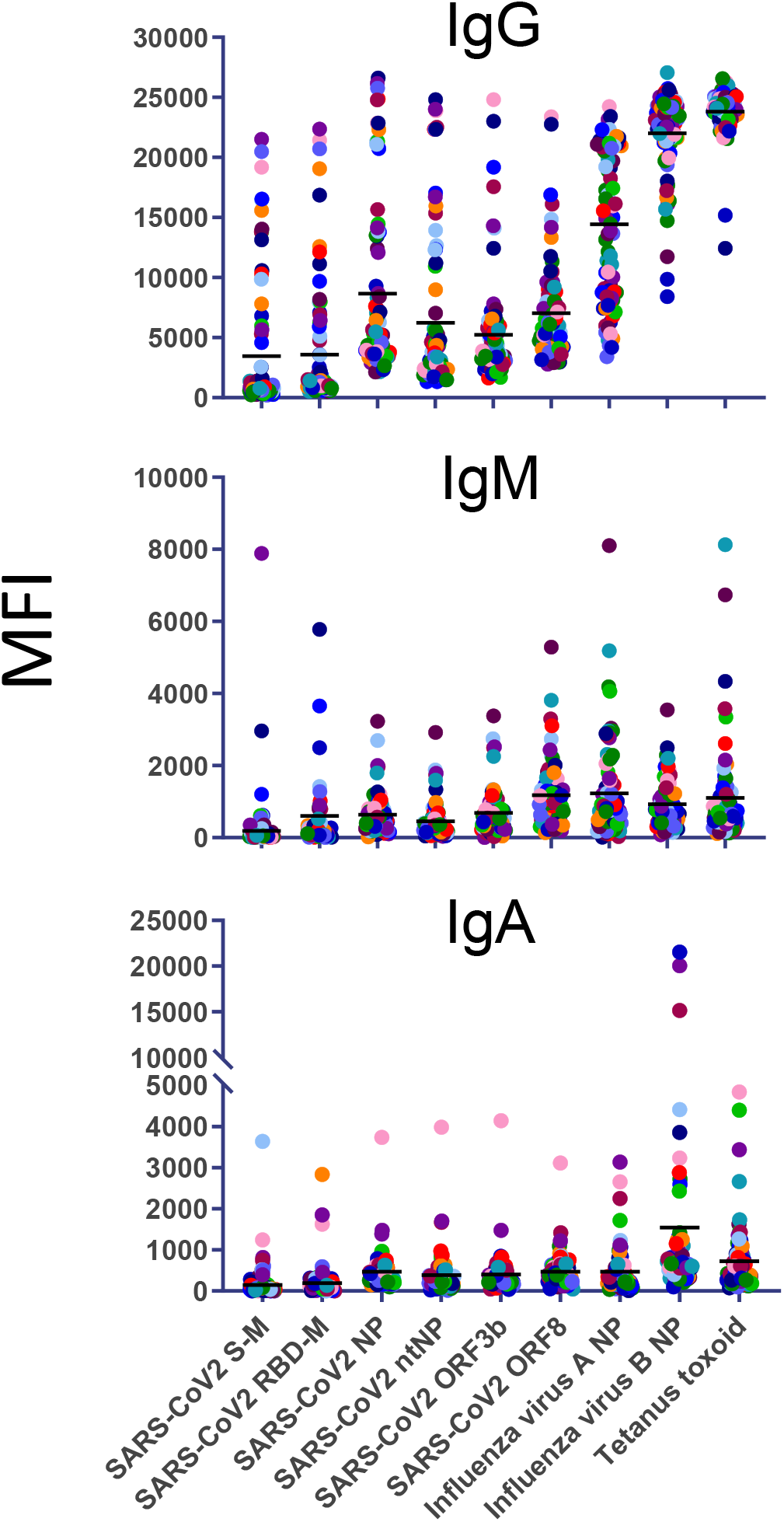
Serum/plasma IgG, IgM and IgA responses to SARS-CoV-2 and influenza and tetanus reference antigens in cohort 2.

Because of the need to accomplish large-scale population-level testing, there has been interest in adapting traditional serologic tests for use with specimen types that can be easily collected in diverse settings where phlebotomy may be impractical. Therefore, we assessed the correlation of antibody levels measured in serum, saliva, and/or dried blood spot (DBS) eluate. As shown in Fig. 4, serum and DBS gave nearly identical results, while saliva showed variability in the specific antigens recognized, as well as lower levels for all Ig isotypes in general.

**Figure 4.**
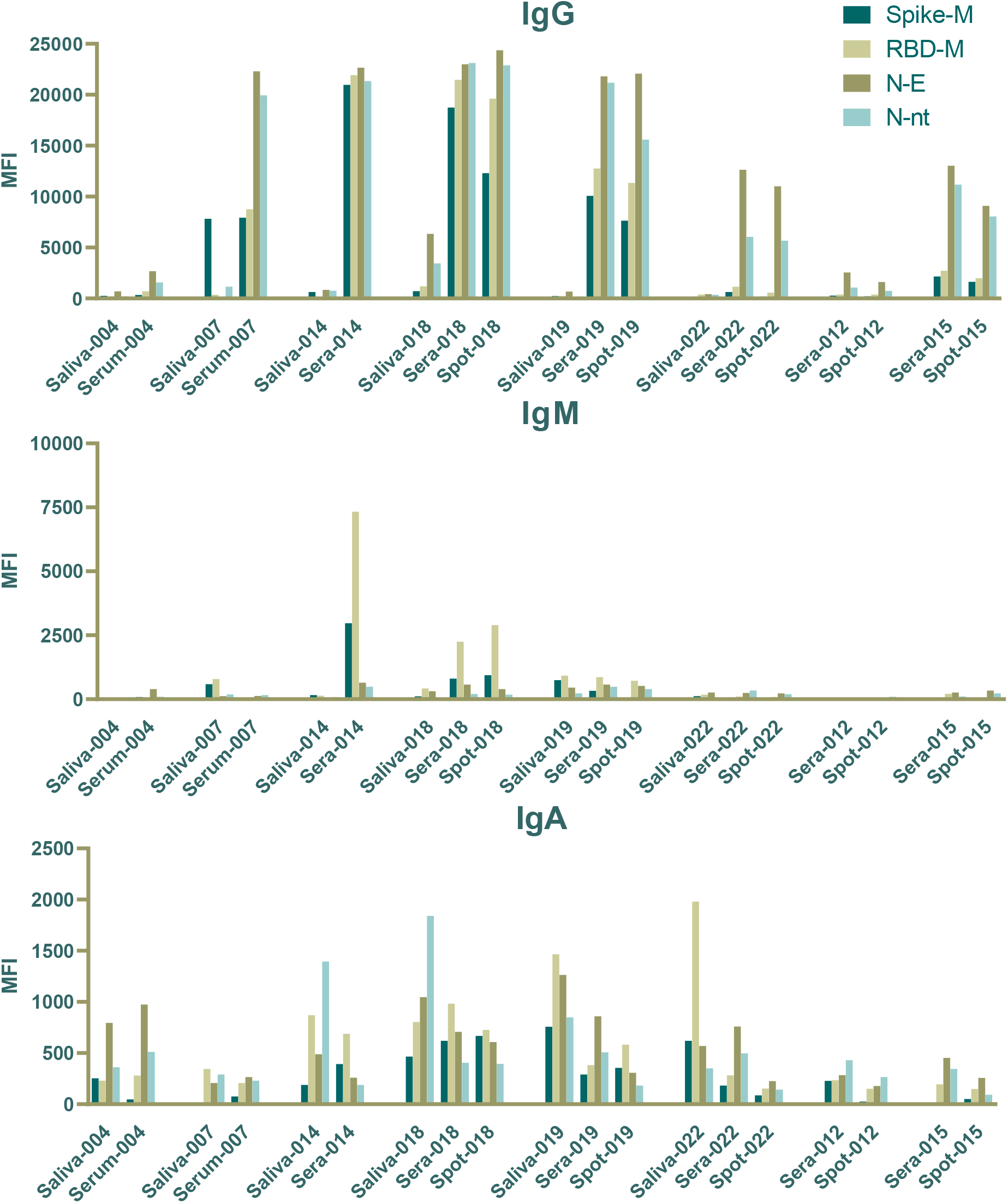
Concordant detection of SARS-CoV-2 antigen-specific of IgG, IgM and IgA in serum and blood spots and differential IgA detection in saliva from a subset of cohort 1 subjects.

Luminex-based assays excel at quantitative monitoring of changes in antibody levels over time (1-7). We analyzed temporal trends in levels of SARS-CoV-2-specific antibody binding for a subset of individuals following COVID-19 symptoms (Fig. 5). Considerable individual variation was observed. Some samples confirmed early generation of IgM, IgG and IgA responses, while others demonstrate the relative stability of IgG responses for >100 days in some cases. Still other cases exhibited increasing IgG levels between 40 and 100 days post symptom onset. The Flu- and TT-specific responses were relatively unchanged across time points.

**Figure 5.**
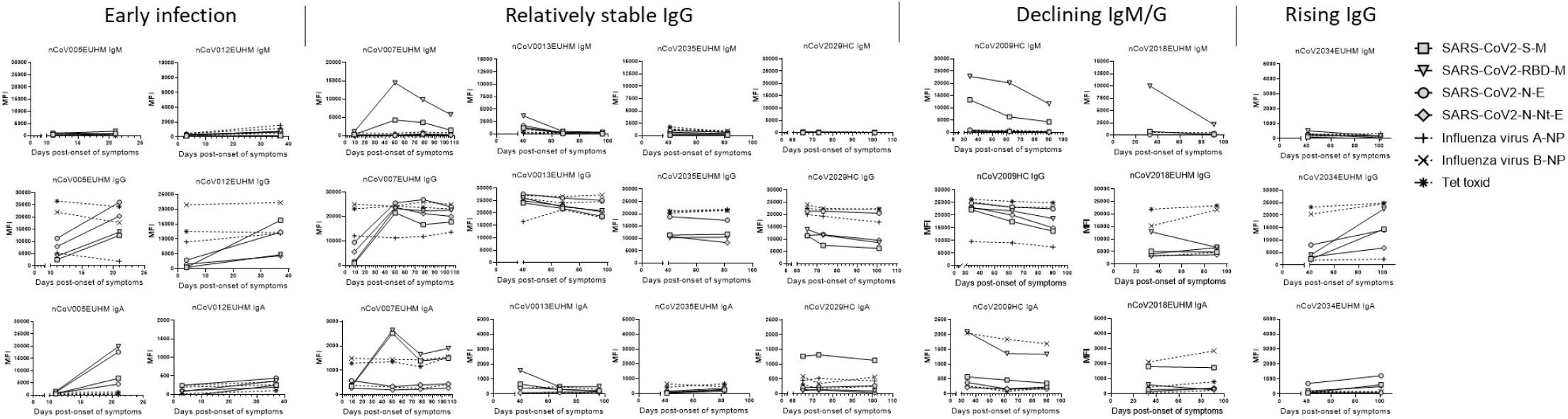
Variation in SARS-CoV-2-specific IgM, IgG and IgA with stable reference antigen-specific antibodies in sera from Cohort 3 subjects collected over time post onset of symptoms.

In addition to humans, SARS-CoV-2, is also known to infect a variety of other species, including non-human primates (9,10) and household pets (11-13). To further evaluate the flexibility of the Luminex-based approach, we tested samples from dogs and cats from households with a confirmed human SARS-CoV-2 infection. Animals were classified as SARS-CoV-2 immune or non-immune based on previously performed testing for SARS-CoV-2 neutralizing antibodies (13). As shown in Fig. 6, modest, to high IgG antibodies could be detected in both dogs and cats in the same assay system using species-specific detecting antibodies.

**Figure 6.**
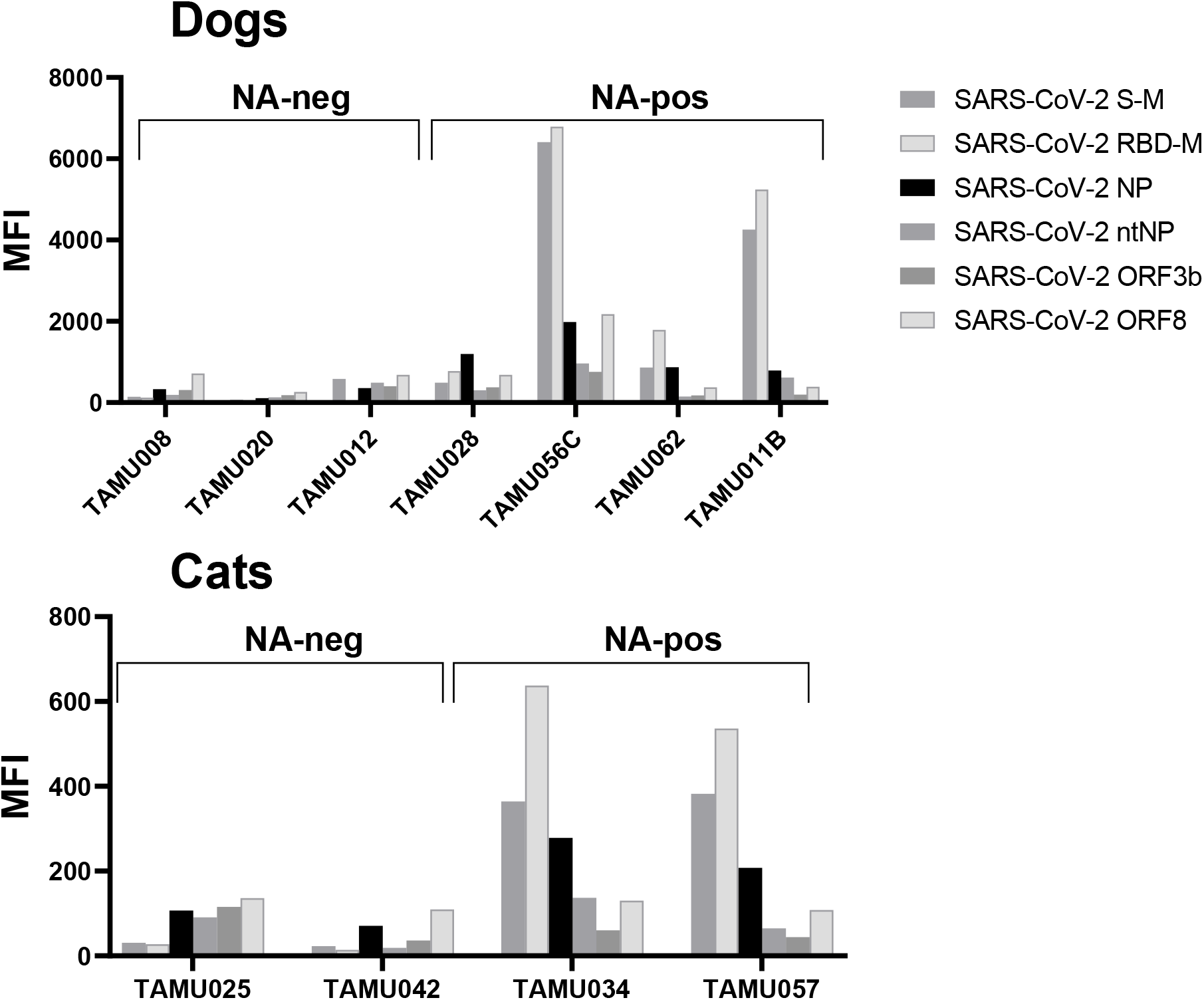
Multiplex antigen detection of SARS-CoV-2-specific antibodies in sera from SARS-CoV-2-neutralizing antibody positive (NA-pos) but not NA-neg dogs and cats.

Recent studies have begun to provide information on the epitope specificity of SARS-CoV-2 Spike-specific antibodies in COVID-19 patients, including regions that are the targets of neutralizing antibodies (14,15). Using these data, we designed and produced 2 new constructs in *E. coli*: Spike-4P contains the 20-mer Spike epitopes aa 550-570, 785-805, 810-830, and 1146-1166 (15) connected by GS amino acid linkers and protein Spike 403-505 contains Spike aa 403-505 which encompassed the region within the RBD recognized by several neutralizing monoclonal antibodies (14). Both proteins were specifically recognized by subjects in cohorts 2 and 3 but not by negative controls, demonstrating the specificity of these responses for epitopes expressed in SARS-CoV-2 infected subjects (Fig. 7A). However, for both antigens, the detection of antibodies was substantially different (either higher or lower) than that detected by the full Spike-M proteins. This differential response to full-length Spike and the hybrid antigens was confirmed in longitudinal specimens, with responses sometimes changing in entirely different directions in the months after infection (Fig. 7B).

**Figure 7.**
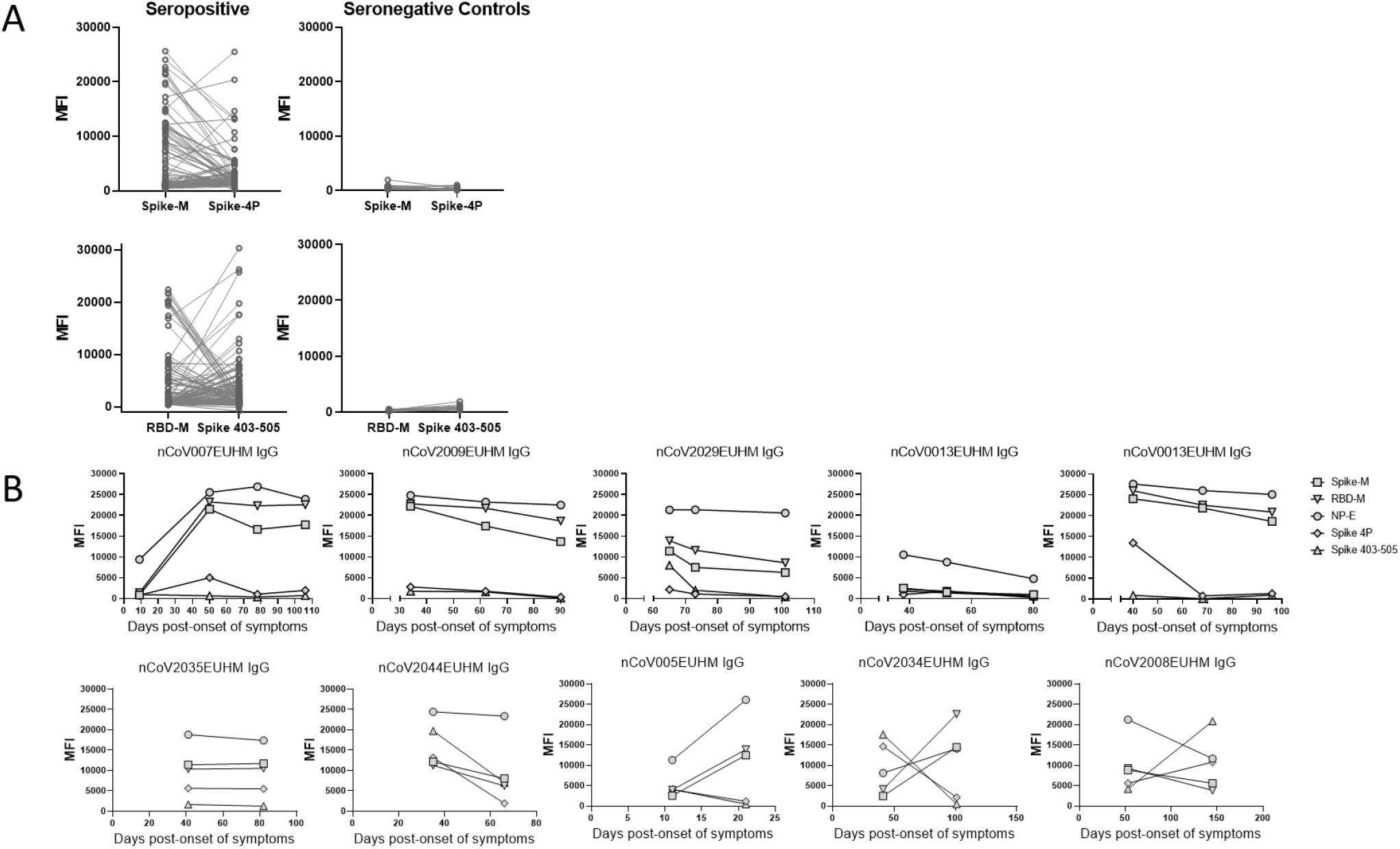
Differential detection of antibodies specific for Spike-M and RBD-M proteins relative to a hybrid protein containing 4 20aa-long epitopes (Spike-4P) or a fragment of the RBD capable of binding neutralizing monoclonal antibodies (Spike 403-505), respectively (A). Matched samples from each Cohort 2 subject or pre-pandemic controls are connected. Longitudinal serum IgG specific for Spike-M, RBD-M, NP-E, Spike-4P and Spike 403-505 in a subset of cohort 3 subjects (B).

## Discussion

The goal of this study was to rapidly develop a serological assay capable of detecting multiple isotypes and antigen specificities of SARS-CoV-2-specific antibodies in various sample types from multiple species, using reagents that can be easily acquired or cheaply produced in most biomedical labs. We knew from previous work in Chagas disease (1-7) that the Luminex multiplexing system met most of these criteria, in addition to having high precision (a result of >50 determinations obtained per sample/antigen determination in this bead-based method), a wide dynamic range, and requiring minimal sample amounts. All of the antigens used can be easily produced and purified in-house and/or resourced commercially. In the former case, a single 150 ml *E. coli* culture provides sufficient protein for >30,000 determinations and up to 100 different antigens can be assessed in each sample using a total of <1 ul of serum or the eluate from a blood spot. Depending on the number of antigens and isotypes assessed, and the sources of such reagents, the cost for each assay can be as low as $5. The *E. coli*-expression plasmids developed in this study are being made available through Addgene to facilitate their broader use and the protocols for protein production and bead conjugation are straightforward. The main limitation in adopting this multiplexing approach is the requirement for access to a Luminex assay reader, although these instruments are becoming increasingly available and can be purchased new for <$30,000 or much less for second-hand instruments.

During the course of this work, many reports of serological diagnostic assays, including those based upon the Luminex format (15-23), have been presented. Rather than developing an additional diagnostic platform, we emphasize here the utility of the Luminex system as a research tool – for monitoring changes in antibody responses over time, detecting antibody responses in different sample matrices and for rapid adaptation and evaluation of new antigen formulations. Using this tool, we reconfirm the wide variation in the generation and maintenance, as well as antigen-specificity of SARS-CoV-2-specific antibodies in different individuals/animals and using multiple sample types, as well as the various patterns of change in these antibody levels over time post-infection. Including multiple SARS-CoV-2 antigens in a single assay not only potentially increases the sensitivity of detection of infection history (17), it also enhances the relative depth of the data on immune response patterns, without the time and sample volume requirements of individual assays. The inclusion of reference antigens such as influenza nucleoproteins and tetanus toxoid provide confidence that the observed changes in SARS-CoV-2-specific responses are not the result of sample quality/storage. These non-SARS antigens, as well as SARS-CoV-2 antigens not part of vaccine regimens could also serve as crucial reference points for tracking the boosting of the SARS-CoV-2-specific responses due to re-exposure or vaccination and serologic surveillance for incident infection in vaccines.

Among the most important observations from these studies was the differential detection of deglycosylated Spike protein as well as linked Spike epitopes and Spike fragments by serum antibodies from previously infected subjects. These results suggest that the failure of Spike protein produced in *E. coli* to bind SARS-CoV-2-specific antibodies was not due to the absence of glycans known to decorate the mammalian-produced proteins, but rather likely to defects in protein folding that were not an issue in the Spike-4P and Spike 403-505 fragments produced in *E. coli*. While it may be interesting to explore the biological relevance of these antibodies recognizing the different Spike formulations and fragments in future studies, these results further reinforce the value of having multiple antigens – including new constructs derived from epitope-discovery efforts – tested in parallel in serological assays. It is noteworthy that for the Spike-4P and Spike 403-505, the time required to go from their identification in published reports to design of the constructs and inclusion of the proteins in assays in our lab, was <40 days (and could have been done even faster if necessary). The association of the changing pattern of responses to these and other SARS-CoV-2 epitopes could be of use in identifying response patterns that associate with risk of infection/reinfection, or protection from severe disease.

Lastly, the ease with which this platform can be used with a variety of sample types, including those that can be self-collected by study subjects, and applied to multiple species with only minor modification, makes it expedient for a wide range of applications and study types. The simultaneous assessment of multiple antigens and the capacity to readily incorporate new or additional antigens is especially useful for novel pathogens such as SARS-CoV-2, where the preferential immunogenic epitopes may not be immediately available or widely known.

## Materials and Methods

### Study Design

Three human sample sets and one domestic animal sample set were used to evaluate the Luminex-based serological assay. Pre-pandemic sera collected prior to July 2019 as part of a study of serological responses in Chagas disease subjects were used as virus-negative controls in some assays.

For human cohort 1, eligible volunteers between the ages of 18 and 65 years old (y.o.), were recruited from residents in Georgia, USA and enrolled with written, informed consent. Exclusion criteria included documented dementia or Alzheimer disease, estimated life expectancy <2 years, and medical treatment causing or diagnosis of an immunocompromising condition. “P” subjects were negative by ELISA for IgG specific for the SARS-CoV-2 RBD region of the Spike protein, and were not tested for viral RNA. All “A” subjects were SARS-CoV-2 RT-PCR as well as ELISA-positive. Cohort 1 subjects 014, 018, and 019 were both ELISA and viral positive and 004, 012, and 022 were both ELISA and viral negative. Subject 007 showed COVID symptoms and tested ELISA positive but was not tested for virus and 015 tested viral positive three months prior to a negative ELISA test.

For human cohorts 2 and 3, serum/plasma samples were provided by collaborators from Emory University from participants in two settings: cohort 2) Samples (n=80) were obtained during May and June, 2020, from healthcare personnel enrolling in a SARS-CoV-2 surveillance study. Samples shared included known seronegatives and a subset of samples enriched for seropositive samples according to previous testing by a SARS-CoV-2 RBD ELISA to detect IgG and cohort 3) PCR-confirmed COVID-19 cases invited to donate specimens at various times during and/or after recovering from their acute illness.

Participants provided verbal and / or written informed consent and provided blood specimens for analysis. Only de-identified serum or plasma including limited metadata (days post-symptom onset [DPSO] and SARS-CoV-2 RT-PCR status [either positive or negative]) were shared for this study.

Previously generated serologic data from cohort 2 was not shared.

Blood was sampled from dogs and cats from central Texas as part of a longitudinal epidemiological investigation of SARS-CoV-2 infection in pets living with at least one human with laboratory confirmed SARS-CoV-2 infection (13). Serum aliquots from dogs and cats that tested positive for neutralizing antibodies and met the USDA case definition were shared for the current study, in addition to aliquots from dogs and cats that tested negative.

### Sample collection

For human subjects, blood samples were collected into ACD (acid, citrate, dextrose), EDTA (ethylenediaminetetraacetic acid) or serum separator tubes (SST). Serum was allowed to clot for at least 20 min at room temperature. ACD and EDTA were stored at 4°C for <48 hours before being heat inactivated at 60°C for 30 minutes, aliquoted and stored at -80°C or -150°C. For a subset of subjects, blood spots and sputum samples (diluted ½ in viral transport medium were collected at the same time as blood draws and stored at room temperature and at -80°C respectively.

From dog and cat subjects, blood samples were collected into clot activator tubes. After centrifugation, serum aliquots were stored at -80°C and tested for the presence of SARS-CoV-2 neutralizing antibodies at the National Veterinary Services Laboratory as previous described (13).

### Protein production

Codon optimized expression constructs in pET30a or pET32a vectors between BamH I and Hind III sites with an in-frame N-terminal 6X-histidine metal affinity tag (GenScript, Piscataway, NJ) were used for recombinant protein production in *E. coli* BL21(DE3) as previous described (Cooley, 2008) (Supplemental Table S1).

Recombinant SARS-CoV-2 full-length Spike and receptor-binding domain (RBD) proteins were produced as described (24,25). Briefly, plasmids encoding the RBD or Spike protein with C-terminal 6xHis tags were used to transfect Expi293 cells and proteins were purified from clarified supernatants by nickel resin chromatography and dialyzed into PBS. Proteins were quantitated, aliquoted, and stored at -80°C. Protein purity and folding were confirmed by western blot (using anti-6xHis detection antibody) and ELISA (using CR3022 mAb), respectively.

For deglycosylation, recombinant Spike protein produced and purified as previously described (26) was deglycosylated using PNGase F (produced as previously described in (27)), at a ratio of 20:1 of Spike to enzyme at 4°C overnight. Deglycosylated Spike trimer was then purified by Superdex G-200 column (GE Healthcare) gel filtration and the size confirmed by SDS PAGE analysis.

### Multiplex Serological Assays

Proteins were attached to Luminex beads and Luminex assays were conducted as described previously (3). Phycoerythrin-labeled secondary antibodies for various species were obtained from Jackson ImmunoResearch anti-human IgG, cat#709-116-149, IgM, cat#706-116-073, and IgA, cat#109-115-011), Rockland Immunochemical (anti-canine Ig cat#RL704-408-002; and Novus biologicals (anti-feline Ig cat#NBP2-60651PEATT594). Data are expressed as raw mean fluorescence intensity (MFI) from a minimum of 50 beads per determination. All the samples were diluted with dilution buffer (PBS, 1 mg/ml casein, 0.5% polyvinyl alcohol (PVA), 0.8% polyvinyl pyrrolidone (PVP) and 3 mg/ml E. coli lysate). Human serum/plasma samples were diluted 1/100, saliva samples were assayed at a final dilution of 1/8 and blood spot samples were eluted as described previously (28). Dog and cat serum samples were diluted 1/50.

### ELISA Serological Assays

Immulon^®^ 4HBX plates (Thermo Fisher Scientific, Waltham, MA, USA) were coated with 100 ng/well of the receptor binding domain (RBD) of the SARS-CoV-2 Spike protein in PBS overnight at 4° C in a humidified chamber. Plates were blocked with blocking buffer containing 2% bovine serum albumin (BSA) Fraction V (Thermo Fisher Scientific, Waltham, MA, USA), 1% gelatin from bovine skin (Sigma-Aldrich, St. Louis, MO, USA) in PBS/0.05% Tween20 (Thermo Fisher Scientific, Waltham, MA, USA) at 37° C for 90 minutes. Serum samples were initially diluted 1:50, and then further serially diluted 1:3 in blocking buffer to generate a 4-point binding curve (1:50, 1:150, 1:450, 1:1350) and incubated overnight in the RBD-coated plates at 4° C in a humidified chamber. Plates were washed 5 times with PBS with 0.05% Tween20 added and IgG antibodies were detected using horseradish peroxidase (HRP)-conjugated goat anti-human IgG detection antibody (Southern Biotech, Birmingham, AL, USA) at a 1:4000 dilution and incubated for 90 minutes at 37° C. Plates were then washed 5 times with PBS with 0.05% Tween20 prior to development with 100μL of 0.1% 2,2’-azino-bis(3-ethylbenzothiazoline-6-sulphonic acid) (ABTS, Bioworld, Dublin, OH, USA) solution with 0.05% H_2_O_2_ for 18 min at 37° C. The reaction was terminated with 50μL of 1% (w/v) sodium dodecyl sulfate (SDS, VWR International, Radnor, PA, USA). Colorimetric absorbance at 414nm was measured using a PowerWaveXS (Biotek, Winooski, VT, USA) plate reader. Each participant serum and controls were run in duplicates and the mean of the two optical density (OD) values were used in downstream analyses.

### Statistical and graphical analysis

Statistical analysis and graphical plots were done in Prism 9 (GraphPad Software, San Diego, CA).

### Study approval

Participation in these studies was voluntary. Sample collection at the University of Georgia (cohort 1) was reviewed and approved by the University of Georgia Institutional Review Board (Protocol # 20202906) and at Emory University was performed under two protocols, which were reviewed by the Emory University Institutional Review Board: IRB#00000505 (cohort 2) and IRB#00022371 (cohort 3). All pet samples were obtained from privately-owned animals in adherence with animal use protocols approved by the Texas A&M University’s Institutional Animal Care and Use Committee (IACUC) and Clinical Research Review Committee (CRRC) on May 14, 2020 (2018-0460 CA). Written consent was received for each pet from the owner. The funding source had no role in sample collection nor decision to submit the paper for publication.

## Data Availability

All data generated or analysed during this study are included in this published article (and its supplementary information files).

## Author Contributions

HS designed the studies, conducted experiments and acquired and analyzed data. DC conducted experiments and provided reagents, DF, KWM, SAH, SMT, NR, SE and TMR provided reagents and edited the manuscript. LW provided reagents. MHC helped design the studies, provided reagents and edited the manuscript. RLT designed the studies, analyzed the data and wrote the manuscript.

## Acknowledgements

We thank the participants who provided blood or sputum specimens that were tested in this study. We thank the faculty and staff team of The Hope Clinic for a dedicated effort in recruiting subjects and collecting specimens and Hannah Hanley and Mitchell Lee for the processing of the serum and saliva samples. We thank Dr. Yerun Zhu and Dr. Daniel Espinoza for assistance with laboratory operations. We thank Italo Zecca, Lisa Auckland, Edward Davila, Gabriel Hamer, Chris Roundy, and Wendy Tang for collecting samples from pets. Mia Torchetti, Melinda Jenkins Moore, and Katie Mozingo provided virus neutralization testing of animal samples. Plasmids for expression of SARS-CoV-2 Spike and RBD proteins, as well as CR3022 monoclonal antibody were generously provided by Dr. Florian Krammer (Icahn School of Medicine at Mount Sinai, produced under NIAID CEIRS contract HHSN272201400008C). Financial support for this work was provided by grant R01AI125738 to RLT from the National Institutes of Health and a University of Georgia Athletic Association endowment to RLT. S.M.T. and T.M.R. are partially supported by NIAID Centers of Excellence for Influenza Research and Surveillance (CEIRS) contract HHSN272201400004C and from the University of Georgia. T.M.R and LW are also supported, in part, by the Georgia Research Alliance as an Eminent Scholar and a Distinguished Investigator, respectively. Additional support for specimen collection and processing of samples in cohort 2 was provided by the Georgia Emerging Infections Program, which was funded through the Centers for Disease Control and Prevention Emerging Infections Program [U50CK000485]. Pet dog and cat samples were collected with funding from the Centers for Disease Control and Prevention RFP 75D 301-20-R-68167. The findings and conclusions in this report are those of the authors and do not necessarily represent the official position of the Centers for Disease Control and Prevention.

---

**Supplemental Table S1.**

| Protein | Description | Expression plasmid | Cloning site | Protein sequence |
| --- | --- | --- | --- | --- |
| SARS-CoV2-ORF3b (S23Q) | Stop codon to glutamine at 23<sup>rd</sup> amino acid | pET-30a(+) | BamHI/HindIII | Spike-E |
| SARS-CoV2-ORF8 |  | pET-30a(+) | BamHI/HindIII |  |
| SARS-CoV2-NP | SARS-CoV2 nucleocapsid protein | pET-30a(+) | BamHI/HindIII |  |
| SARS-CoV2-NP-Nt(47-173) | SARS-CoV2 nucleocapsid protein N-terminal | pET-30a(+) | BamHI/HindIII |  |
| FluA-NP | Influenza A virus nucleocapsid protein | pET-30a(+) | BamHI/HindIII |  |
| FluB-NP | Influenza B virus nucleocapsid protein | pET-30a(+) | BamHI/HindIII |  |
| Spike-S-4P | SARS-CoV2 spike protein site 550-570, 785-805, 810-830 and 1146-1166 | pET-32a(+) | BamHI/HindIII |  |
| Spike-403-505 | SARS-CoV2 spike protein site 403-505 | pET-32a(+) | BamHI/HindIII |  |
| Spike-E | SARS-CoV2 Spike protein | pET-30a(+) | BamHI/HindIII |  |

## Nucleotide sequences

### SARS-CoV-2 Spike-E

ATGTTCGTTTTCCTGGTTCTGCTGCCGCTGGTTAGCAGCCAATGCGTGAATCTGACCACCCGCACCCAACTGCCGCCGGCGTACACCAACAGCTTCACCCGTGGTGTTTACTATCCGGACAAAGTTTTTCGTAGCAGCGTGCTGCACAGCACCCAGGACCTGTTCCTGCCGTTCTTTAGCAACGTTACCTGGTTCCACGCGATCCACGTGAGCGGCACCAACGGCACCAAGCGTTTCGACAACCCGGTGCTGCCGTTTAACGATGGTGTTTACTTCGCGAGCACCGAGAAGAGCAACATCATTCGTGGTTGGATTTTTGGCACCACCCTGGACAGCAAAACCCAGAGCCTGCTGATCGTTAACAACGCGACCAACGTGGTTATTAAGGTGTGCGAATTCCAATTCTGCAACGATCCGTTCCTGGGCGTTTACTATCACAAGAACAACAAAAGCTGGATGGAGAGCGAATTTCGTGTTTATAGCAGCGCGAACAACTGCACCTTTGAGTACGTGAGCCAGCCGTTCCTGATGGACCTGGAAGGCAAGCAAGGCAACTTCAAAAACCTGCGTGAGTTCGTGTTCAAGAACATCGATGGTTACTTCAAGATCTACAGCAAGCACACCCCGATCAACCTGGTTCGTGACCTGCCGCAGGGTTTTAGCGCGCTGGAGCCGCTGGTTGACCTGCCGATCGGCATTAACATCACCCGTTTTCAAACCCTGCTGGCGCTGCACCGTAGCTACCTGACCCCGGGTGACAGCAGCAGCGGTTGGACCGCGGGTGCGGCGGCGTACTATGTTGGTTATCTGCAGCCGCGTACCTTCCTGCTGAAATACAACGAAAACGGCACCATCACCGACGCGGTTGATTGCGCGCTGGATCCGCTGAGCGAAACCAAGTGCACCCTGAAAAGCTTTACCGTGGAGAAGGGTATTTATCAGACCAGCAACTTCCGTGTGCAACCGACCGAAAGCATTGTTCGTTTTCCGAACATCACCAACCTGTGCCCGTTTGGCGAGGTTTTCAACGCGACCCGTTTCGCGAGCGTGTACGCGTGGAACCGTAAACGTATCAGCAACTGCGTTGCGGACTATAGCGTGCTGTACAACAGCGCGAGCTTCAGCACCTTTAAGTGCTATGGTGTGAGCCCGACCAAACTGAACGATCTGTGCTTTACCAACGTTTACGCGGATAGCTTCGTGATTCGTGGCGACGAGGTTCGTCAGATCGCGCCGGGTCAAACCGGCAAGATTGCGGACTACAACTATAAACTGCCGGACGATTTCACCGGTTGCGTTATCGCGTGGAACAGCAACAACCTGGATAGCAAAGTGGGTGGCAACTACAACTATCTGTACCGTCTGTTTCGTAAGAGCAACCTGAAACCGTTCGAGCGTGACATTAGCACCGAAATCTACCAGGCGGGTAGCACCCCGTGCAACGGTGTTGAGGGCTTTAACTGCTATTTCCCGCTGCAGAGCTACGGCTTCCAACCGACCAACGGTGTGGGCTATCAACCGTACCGTGTGGTTGTGCTGAGCTTTGAACTGCTGCATGCGCCGGCGACCGTTTGCGGTCCGAAGAAAAGCACCAACCTGGTTAAGAACAAATGCGTGAACTTCAACTTTAACGGCCTGACCGGCACCGGTGTGCTGACCGAGAGCAACAAGAAATTCCTGCCGTTTCAGCAATTCGGTCGTGACATTGCGGATACCACCGATGCGGTGCGTGACCCGCAGACCCTGGAAATTCTGGATATCACCCCGTGCAGCTTCGGTGGCGTTAGCGTGATCACCCCGGGCACCAACACCAGCAACCAGGTTGCGGTGCTGTATCAAGACGTTAACTGCACCGAGGTTCCGGTGGCGATTCACGCGGATCAGCTGACCCCGACCTGGCGTGTGTACAGCACCGGTAGCAACGTTTTCCAAACCCGTGCGGGTTGCCTGATTGGTGCGGAGCATGTGAACAACAGCTATGAATGCGACATTCCGATCGGTGCGGGCATTTGCGCGAGCTACCAGACCCAAACCAACAGCCCGGGTAGCGCGAGCAGCGTTGCGAGCCAGAGCATCATTGCGTATACCATGAGCCTGGGCGCGGAAAACAGCGTGGCGTACAGCAACAACAGCATTGCGATCCCGACCAACTTCACCATTAGCGTGACCACCGAGATCCTGCCGGTTAGCATGACCAAAACCAGCGTGGACTGCACCATGTATATCTGCGGTGATAGCACCGAATGCAGCAACCTGCTGCTGCAGTACGGTAGCTTTTGCACCCAACTGAACCGTGCGCTGACCGGCATTGCGGTGGAGCAGGATAAAAACACCCAAGAAGTTTTCGCGCAGGTGAAGCAAATTTACAAAACCCCGCCGATCAAGGACTTTGGTGGCTTCAACTTTAGCCAGATCCTGCCGGATCCGAGCAAGCCGAGCAAACGTAGCTTTATTGAGGACCTGCTGTTCAACAAGGTTACCCTGGCGGATGCGGGTTTCATCAAACAGTATGGTG ATTGCCTGGGCGACATTGCGGCGCGTGACCTGATCTGCGCGCAAAAGTTTAACGGCCTGACCGTGCTGCCGCCGCTGCTGACCGATGAAATGATTGCGCAGTACACCAGCGCGCTGCTGGCGGGCACCATTACCAGCGGTTGGACCTTTGGTGCGGGTGCGGCGCTGCAGATCCCGTTTGCGATG CAAATGGCGTATCGTTTCAACGGTATTGGCGTTACCCAAAACGTGCTGTACGAGAACCAGAAGCTGATCGCGAACCAATTTAACAGCGCGATTGGTAAAATCCAGGATAGCCTGAGCAGCACCGCGAGCGCGCTGGGTAAACTGCAAGATGTTGTGAACCAGAACGCGCAAGCGCTGAACACCCTGGTTAAGCAGCTGAGCAGCAACTTCGGTGCGATTAGCAGCGTGCTGAACGATATTCTGAGCCGTCTGGACCCGCCGGAGGCGGAAGTTCAAATTGACCGTCTGATCACCGGCCGTCTGCAGAGCCTGCAAACCTATGTGACCCAGCAACTGATTCGTGCGGCGGAAATTCGTGCGAGCGCGAACCTGGCGGCGACCAAAATGAGCGAGTGCGTTCTGGGTCAGAGCAAGCGTGTGGACTTTTGCGGTAAAGGCTATCACCTGATGAGCTTCCCGCAGAGCGCGCCGCACGGTGTTGTGTTTCTGCACGTTACCTACGTGCCGGCGCAAGAAAAGAACTTTACCACCGCGCCGGCGATCTGCCATGATGGTAAAGCGCACTTTCCGCGTGAAGGTGTTTTCGTGAGCAACGGCACCCACTGGTTTGTTACCCAGCGTAACTTCTACGAGCCGCAAATCATTACCACCGACAACACCTTCGTGAGCGGTAACTGCGATGTTGTGATTGGCATCGTTAACAACACCGTGTATGATCCGCTGCAGCCGGAGCTGGACAGCTTTAAAGAGGAGCTGGATAAGTACTTCAAAAACCACACCAGCCCGGACGTTGATCTGGGTGACATTAGCGGCATCAACGCGAGCGTTGTGAACATTCAAAAGGAGATCGACCGTCTGAACGAAGTGGCGAAAAACCTGAACGAAAGCCTGATCGATCTGCAGGAGCTGGGTAAATATGAACAAGGTAGCGGCTACATTCCGGAGGCGCCGCGTGACGGTCAGGCGTATGTGCGTAAGGATGGCGAGTGGGTTCTGCTGAGCACCTTTCTGGGTCGTAGCCTGGAAGTTCTGTTTCAAGGTCCGGGT

### SARS-CoV2-ORF3b (S23Q)

ATGATGCCGACCATCTTCTTTGCGGGTATCCTGATTGTGACCACCATCGTTTACCTGACCATTGTGCAGCTGCTGCAACTGAGCCTGCTGCAGGTGATGGCGCAGCAAGTTCTGTTCCTGAACATGACCACCCGTCTGGTGGTTATTCTGAAGAACGGCAACCTGGAGTAA

### SARS-CoV2-ORF8

ATGAAGTTCCTGGTGTTTCTGGGTATCATTACCACCGTTGCGGCGTTCCACCAGGAGTGCAGCCTGCAAAGCTGCACCCAGCACCAACCGTACGTGGTTGACGATCCGTGCCCGATCCACTTTTACAGCAAGTGGTATATTCGTGTGGGTGCGCGTAAAAGCGCGCCGCTGATCGAGCTGTGCGTTGATGAAGCGGGTAGCAAGAGCCCGATTCAGTACATCGACATTGGCAACTATACCGTGAGCTGCCTGCCGTTCACCATCAACTGCCAAGAACCGAAACTGGGTAGCCTGGTGGTTCGTTGCAGCTTCTACGAGGACTTTCTGGAATATCACGATGTTCGTGTGGTTCTGGACTTTATTTAA

### SARS-CoV2-NP

ATGAGCGACAATGGTCCGCAGAATCAGCGTAATGCGCCGCGTATCACCTTTGGTGGTCCGAGCGACAGCACCGGCAGCAATCAGAACGGTGAACGTAGCGGTGCGCGTAGCAAGCAGCGTCGTCCGCAAGGTCTGCCGAACAACACCGCGAGCTGGTTCACCGCGCTGACCCAGCACGGCAAGGAAGACCTGAAATTTCCGCGTGGTCAAGGCGTGCCGATCAACACCAACAGCAGCCCGGACGATCAGATTGGTTACTATCGTCGTGCGACCCGTCGTATCCGTGGTGGCGACGGCAAGATGAAAGATCTGAGCCCGCGTTGGTACTTCTACTATCTGGGTACCGGTCCGGAGGCGGGTCTGCCGTATGGCGCGAACAAGGACGGTATCATTTGGGTGGCGACCGAAGGTGCGCTGAACACCCCGAAAGATCACATTGGTACCCGTAACCCGGCGAACAACGCGGCGATCGTTCTGCAACTGCCGCAAGGTACCACCCTGCCGAAAGGTTTTTATGCGGAGGGCAGCCGTGGTGGCAGCCAGGCGAGCAGCCGTAGCAGCAGCCGTAGCCGTAACAGCAGCCGTAACAGCACCCCGGGTAGCAGCCGTGGTACCAGCCCGGCGCGTATGGCGGGTAACGGTGGCGACGCGGCGCTGGCGCTGCTGCTGCTGGATCGTCTGAACCAGCTGGAGAGCAAGATGAGCGGTAAAGGCCAGCAACAGCAAGGCCAAACCGTGACCAAGAAAAGCGCGGCGGAAGCGAGCAAGAAACCGCGTCAGAAGCGTACCGCGACCAAAGCGTACAACGTTACCCAAGCGTTCGGTCGTCGTGGTCCGGAGCAGACCCAGGGTAACTTTGGCGACCAGGAACTGATTCGTCAAGGTACCGATTATAAGCACTGGCCGCAAATTGCGCAGTTTGCGCCGAGCGCGAGCGCGTTCTTTGGTATGAGCCGTATTGGCATGGAAGTGACCCCGAGCGGTACCTGGCTGACCTACACCGCGGCGATCAAGCTGGACGATAAAGACCCGAACTTCAAAGATCAGGTTATCCTGCTGAACAAGCACATTGACGCGTATAAAACCTTTCCGCCGACCGAGCCGAAGAAAGACAAGAAAAAGAAAGCGGATGAAACCCAAGCGCTGCCGCAGCGTCAAAAGAAACAGCAAACCGTTACCCTGCTGCCGGCGGCGGATCTGGACGATTTTAGCAAGCAACTGCAACAGAGCATGAGCAGCGCGGACAGCACCCAGGCGGGCGGCGGCGGTAGC

### SARS-CoV2-NP-Nt (47-173)

ATGAACAACACCGCGAGCTGGTTCACCGCGCTGACCCAGCACGGCAAGGAGGACCTGAAATTTCCGCGTGGTCAAGGCGTGCCGATCAACACCAACAGCAGCCCGGACGATCAGATTGGTTACTATCGTCGTGCGACCCGTCGTATCCGTGGTGGTGACGGCAAGATGAAAGATCTGAGCCCGCGTTGGTACTTCTACTATCTGGGTACCGGTCCGGAGGCGGGTCTGCCGTATGGCGCGAACAAGGACGGTATCATTTGGGTGGCGACCGAAGGTGCGCTGAACACCCCGAAAGATCACATTGGTACCCGTAACCCGGCGAACAACGCGGCGATCGTTCTGCAGCTGCCGCAAGGTACCACCCTGCCGAAGGGCTTTTACGCG

### FluB-NP

ATGAGCAACATGGACATTGATGGTATGAACACCGGCACCATTGACAAGACCCCGGAGGAAATCACCAGCGGTACCAGCGGTACCACCCGTCCGATCATTCGTCCGGCGACCCTGGCGCCGCCGAGCAACAAGCGTACCCGTAACCCGAGCCCGGAGCGTGCGACCACCAGCAGCGAAGACGATGTTGGTCGTAAAGCGCAGAAGAAACAAACCCCGACCGAGATCAAGAAAAGCGTGTACAACATGGTGGTTAAGCTGGGTGAATTCTATAACCAG ATGATGGTTAAAGCGGGCCTGAACGACGATATGGAGCGTAACCTGATTCAAAACGCGCACGCGGTGGAACGTATCCTGCTGGCGGCGACCGACGATAAGAAAACCGAGTTTCAGAAGAAAAAGAACGCGCGTGACGTGAAAGAAGGCAAGGAGGAAATTGATCACAACAAGACCGGTGGCACC TTCTACAAGATGGTTCGTGACGATAAAACCATCTATTTTAGCCCGATCCGTATTACCTTCCTGAAAGAGGAAGTGAAGACCATGTACAAAACCACCATGGGCAGCGACGGTTTTAGCGGCCTGAACCACATCATGATTGGCCACAGCCAGATGAACGACGTGTGCTTCCAACGTAGCAAAGCGCTGAAGCGTGTTGGTCTGGACCCGAGCCTGATTAGCACCTTTGCGGGCAGCACCGTTCCGCGTCGTAGCGGTGCGACCGGTGTGGCGATCAAAGGTGGCGGTACCCTGGTTGCGGAAGCGATTCGTTTCATCGGTCGTGCGATGGCGGACCGTGGCCTGCTGCGTGATATTAAAGCGAAGACCGCGTATGAGAAGATCCTGCTGAACCTGAAAAACAAGTGCAGCGCGCCGCAGCAAAAAGCGCTGGTTGATCAGGTGATCGGTAGCCGTAACCCGGGCATTGCGGACATCGAAGATCTGACCCTGCTGGCGCGTAGCATGGTGGTTGTGCGTCCGAGCGTTGCGAGCAAGGTTGTGCTGCCGATCAGCATTTACGCGAAAATTCCGCAGCTGGGTTTTAACGTTGAGGAATACAGCATGGTGGGCTATGAGGCGATGGCGCTGTATAACATGGCGACCCCGGTTAGCATCCTGCGTATGGGTGACGATGCGAAAGACAAGAGCCAACTGTTCTTTATGAGCTGCTTCGGCGCGGCGTACGAGGATCTGCGTGTGCTGAGCGCGCTGACCGGTACCGAATTTAAACCGCGTAGCGCGCTGAAATGCAAGGGCTTCCACGTTCCGGCGAAGGAGCAGGTGGAAGGCATGGGTGCGGCGCTGATGAGCATTAAACTGCAGTTTTGGGCGCCGATGACCCGTAGCGGTGGTAACGAGGCGGGTGGTGACGGTGGTAGCGGTCAGATTAGCTGCAGCCCGGTTTTCGCGGTGGAACGTCCGATCGCGCTGAGCAAGCAAGCGGTGCGTCGTATGCTGAGCATGAACATCGAGGGTCGTGACGCGGATGTTAAGGGCAACCTGCTGAAAATGATGAACGACAGCATGGCGAAAAAGACCAGCGGTAACGCGTTTATTGGCAAAAAGATGTTCCAGATCAGCGATAAAAACAAGACCAACCCGATCGAGATTCCGATCAAACAAACCATCCCGAACTTCTTTTTCGGTCGTGACACCGCGGAAGATTATGACGATCTGGAT

### FluA-NP

ATGGCGAGCCAGGGTACCAAGCGTAGCTACGAGCAAATGGAAACCGGTGGCGAGCGTCAGGACACCACCGAAATCCGTGCGAGCGTGGGTCGTATGATCGGTGGCATTGGCCGTTTCTACATTCAAATGTGCACCGAGCTGAAACTGAGCGACTATGATGGTCGTCTGATCCAGAACAGCATCACCATTGAGCGTATGGTTCTGAGCGCGTTTGACGAACGTCGTAACAAGTATCTGGAGGAACACCCGAGCGCGGGTAAAGATCCGAAGAAAACCGGTGGCCCGATCTACCGTCGTATTGACGGCAAGTGGACCCGTGAGCTGATCCTGTATGATAAAGAGGAAATTCGTCGTGTGTGGCGTCAAGCGAACAACGGTGAAGACGCGACCGCGGGCCTGACCCACATCATGATTTGGCACAGCAACCTGAACGACGCGACCTACCAACGTACCCGTGCGCTGGTTCGTACCGGTATGGACCCGCGTATGTGCAGCCTGATGCAGGGCAGCACCCTGCCGCGTCGTAGCGGTGCGGCGGGTGCGGCGGTGAAGGGTGTTGGCACCATCGCGATGGAGCTGATCCGTATGATTAAACGTGGTATTAACGATCGTAACTTCTGGCGTGGCGAGAACGGCCGTCGTACCCGTGTGGCGTATGAACGTATGTGCAACATCCTGAAGGGCAAATTTCAGACCGCGGCGCAACGTGCGATGATGGACCAGGTTCGTGAAAGCCGTAACCCGGGTAACGCGGAGATCGAAGATCTGATTTTCCTGGCGCGTAGCGCGCTGATTCTGCGTGGCAGCGTGGCGCACAAGAGCTGCCTGCCGGCGTGCGTGTACGGTCTGGCGGTTGCGAGCGGTCACGACTTCGAGCGTGAAGGTTATAGCCTGGTTGGCATCGATCCGTTTAAGCTGCTGCAGAACAGCCAAGTGGTTAGCCTGATGCGTCCGAACGAGAACCCGGCGCACAAAAGCCAACTGGTGTGGATGGCGTGCCACAGCGCGGCGTTCGAAGACCTGCGTGTGAGCAGCTTTATCCGTGGTAAGAAAGTTATTCCGCGTGGCAAGCTGAGCACCCGTGGCGTGCAGATCGCGAGCAACGAGAACGTTGAAACCATGGATAGCAACACCCTGGAGCTGCGTAGCCGTTACTGGGCGATTCGTACCCGTAGCGGTGGCAACACCAACCAGCAAAAAGCGAGCGCGGGTCAGATCAGCGTGCAACCGACCTTCAGCGTTCAACGTAACCTGCCGTTTGAACGTGCGACCGTGATGGCGGCGTTCAGCGGTAACAACGAGGGCCGTACCAGCGACATGCGTACCGAAGTTATTCGTATGATGGAGAGCGCGAAGCCGGAAGATCTGAGCTTCCAGGGTCGTGGCGTGTTTGAGCTGAGCGACGAAAAAGCGACCAACCCGATCGTTCCGAGCTTTGATATGAGCAACGAAGGTAGCTACTTCTTTGGCGACAACGCGGAGGAATATGATAAC

### Spike-4P

ATGAAAAAGACGGGGACAGGAGTACTAACTGAGAGCAATAAAAAATTCCTGCCGTTTCAGCAATTTGGTCGCGATATTGCAGGCTCTATGAAGAAGTTCGCTCAGGTTAAACAAATCTATAAAACCCCGCCTATTAAAGACTTCGGCGGTTTTAACTTCAGCCAGATCGGCTCTATGAAGAAGCCAGATCCGTCGAAGCCGTCCAAACGTAGCTTTATCGAAGATCTGCTGTTTAACAAGGTGACGCTGGCGGATGGTTCCATGAAGAAGTACGACCCGCTCCAACCGGAATTGGACAGCTTCAAAGAGGAGTTGGACAAATACTTCAAGAACCACACCAGC

### Spike-403-505

ATGGGAGATGAAGTAAGGCAAATAGCTCCCGGCCAGACCGGTAAAATCGCTGATTATAACTACAAGCTGCCGGATGACTTTACCGGCTGCGTGATTGCGTGGAATTCCAACAACTTGGACAGCAAGGTGGGCGGTAATTACAATTACCTGTACCGCCTGTTTCGTAAATCTAACCTGAAGCCGTTTGAGCGTGACATCTCGACCGAAATTTATCAGGCAGGTAGCACTCCGTGTAATGGCGTTGAGGGCTTCAACTGCTACTTCCCGTTGCAAAGCTATGG TTTCCAGCCAACGAACGGTGTTGGTTATCAA

